# Expanding the Russian allele frequency reference via cross-laboratory data integration: insights from 7,452 exome samples

**DOI:** 10.1101/2021.11.02.21265801

**Authors:** Yury A. Barbitoff, Darya N. Khmelkova, Ekaterina A. Pomerantseva, Aleksandr V. Slepchenkov, Nikita A. Zubashenko, Irina V. Mironova, Vladimir S. Kaimonov, Dmitrii E. Polev, Victoria V. Tsay, Andrey S. Glotov, Mikhail V. Aseev, Sergey G. Scherbak, Oleg S. Glotov, Arthur A. Isaev, Alexander V. Predeus

## Abstract

Population allele frequency is crucially important for accurate interpretation of known and novel variants in medical genetics. Recently, several large allele frequency databases, such as Genome Aggregation Database (gnomAD), have been created to serve as a global reference for such studies. However, frequencies of many rare alleles vary dramatically between populations, and population-specific allele frequency is often more informative than the global one. Many countries and regions, including Russia, remain poorly studied from the genetic perspective. Here, we report the first successful attempt to integrate genetic information between major medical genetic laboratories in Russia. We construct an open, large-scale reference set of genetic variants by analyzing 7,492 exome samples collected in two major Russian cities of Moscow and St. Petersburg. An approximately tenfold increase in sample size compared to previous studies allowed us to identify genetically distinct clusters of individuals within an admixed population of Russia. We highlight 47 known pathogenic variants that are overrepresented in Russia compared to other European countries. We also identify several dozen high-impact variants that are present in healthy donors despite either being annotated as pathogenic in ClinVar or falling within genes associated with autosomal dominant disorders. The constructed database of genetic variant frequencies in Russia has been made available to the medical genetics community through a variant browser available at http://ruseq.ru.

## Introduction

Next-generation sequencing (NGS) has become a *de facto* standard tool in molecular diagnostics of Mendelian disorders. The advances in the field have been drastic; for example, whole genome sequencing (WGS) screening of all newborns is currently deployed in some countries (https://www.genomicsengland.co.uk/public-dialogue-genomics-newborn-screening/). However, despite rapid introduction of NGS into research and clinical practice, currently only about 42% of all patients with suspected genetic pathology receive a definitive molecular diagnosis from the trio-based WGS analysis (reviewed in (Wright, FitzPatrick, and Firth 2018)). There can be many explanations for the less-than-perfect diagnostic record of NGS-based approaches. However, most researchers agree that our ability to predict the effects of individual variants on human health is currently very limited (Biesecker and Green 2014), and is far lower than our capacity to detect these variants.

One of the most significant advances of our ability to assess variant effects came from global resequencing projects such as 1000 Genomes project (Auton et al. 2015), the Genome Aggregation Database (gnomAD), or National Heart, Blood, and Lung Institute (NHLBI) TopMed (Bick et al. 2020). A simple argument that many variants previously listed as pathogenic are found in healthy individuals in too high a frequency to cause a Mendelian disorder has become the most powerful tool of reducing false positive variant-phenotype associations (Lek et al. 2016). To this end, the information about the population allele frequency (AF) is currently broadly used for variant interpretation in clinical practice. All of the modern variant interpretation strategies and guidelines, such as the American College of Medical Genetics and Genomics (ACMG) guidelines (Richards et al. 2015), Russian variant interpretation guidelines (Ryzhkova et al. 2018), or the Sherloc guidelines (Nykamp et al. 2017) use AF in healthy populations to classify variant effects, which becomes especially critical for autosomal dominant (AD) diseases.

While global allele frequency databases remain widely useful, the additional value of population-specific reference databases was recently highlighted. Even in the original ExAC publication, Lek et al. (Lek et al. 2016) have shown that filtering of candidate variants using the maximum allele frequency across populations substantially decreases the number of potentially disease-causing variants observed in an individual exome sample. Recently, a substantial effort of the genomic community has been directed at the creation of more diverse and inclusive populational reference as well as resources covering diverse ethnic and racial groups (e.g., (Wong et al. 2020); for review see (Martin et al. 2018)). In many countries, nation-wide sequencing projects have been conducted, including Genome of the Netherlands (GoNL, (Boomsma et al. 2014)) or the Han Genome Database (PGG.*Han*, (Gao et al. 2020)).

The Russian population, representing over 160 nationalities and many unique sub-populations, remains one of the biggest white spots on the global map of human genomic diversity (Oleksyk, Brukhin, and O’Brien 2015). Several previous studies focused on investigating the genome-wide variation of the Russian population. These include the pilot phase of the Genomes Russia project (Zhernakova et al. 2020), an exome-based study of monogenic disease prevalence in 694 patients (Barbitoff et al. 2019), and a targeted sequencing study of 242 known disease genes in 1,658 healthy individuals from the Ivanovo region (Ramensky et al. 2021). In these works, several important aspects of the genome variation in Russian patients have been pinpointed. All of these studies, however, lack in the comprehensiveness of the analysis due to either low sample size (such as in the Genomes Russia project) or narrow set of analyzed genes (Ramensky et al. 2021).

Centralized creation of a population genomic reference could benefit from funding security and uniform approaches to sequencing and analysis; at the same time, it is associated with numerous logistical and other difficulties. Most population references were de facto obtained using the aggregation of exome and genome data across multiple sequencing centers. The success of ExAC and gnomAD has proven the efficiency and scalability of this approach (Lek et al. 2016; Karczewski et al. 2020). At the same time, such integration is a challenging task due to differences in both sequencing approaches and data analysis methods used in different laboratories.

In this study, we report the first successful integration of genetic variation data across three major Russian genetic laboratories. To enhance the reproducibility of the analysis and allow a secure and confidential collaboration between the sequencing centers, we developed a portable and reproducible computational pipeline that can be used locally at any future participant laboratory, and subsequently integrated into the global reference using a pre-set analytical and legal framework. We believe that our approach allows for resource- and time-efficient aggregation of data between sequencing centers and will greatly facilitate the construction of a reliable allele frequency reference for the Russian or understudied populations.

## Materials and methods

### Samples and ethics statement

The study comprised groups of patients that were subjected to NGS-based assays for molecular diagnostics and/or genetic screening at three major private centers located in Moscow (Genetico Ltd., denoted “Lab 1”) and St. Petersburg (CerbaLab Ltd. and City Hospital No. 40, denoted “Lab 2” and “Lab 3”, respectively). As detailed below, the dataset contained both healthy and diseased individuals, with healthy donors accounting for 26.1% (1,945/7,452) of all study participants. The “diseased” subgroup included various Mendelian or likely Mendelian and non-Mendelian phenotypes (with the majority of samples having neurological and neuro-muscular disorders (see Supplementary Figure S1)); the “healthy” subgroup comprised healthy donors who were sequenced for carrier screening purposes or patients with multifactorial pathologies, such as obesity or type 2 diabetes. Nearly all patients were residents of the Russian Federation or republics of the former USSR. All participants signed informed consent for studies and processing of personal data, including medical history data. The study was performed in accordance with the Declaration of Helsinki.

### Exome sequencing

DNA for sequencing was extracted either from peripheral blood samples (in the majority of cases) or from tissue samples in FFPE blocks according to the standard protocols. Sequencing was performed using either whole exome (5,418 samples; 72.7%) or clinical exome (2,034 samples, 27.3%) capture kits. The following capture kits were used: Agilent SureSelect Human all exon V7, Agilent SureSelect Human all exon V6 + UTR, TruSeq DNA Exome (Illumina) with the xGen® Exome Research Panel v1.0 (IDT) exome capture solution, Nimblegen (Roche) SeqCap EZ MedExome, Illumina Nextera RapidCapture, Illumina TruSeq DNA Exome, Nimblegen (Roche) Inherited Disease Panel (IDP) v2, and Illumina TruSight One. Exome libraries were prepared as described previously (Barbitoff et al. 2019; 2020). All libraries were sequenced using Illumina HiSeq 2500/4000, Illumina NovaSeq 6000, MiSeq, or MGISEQ 2000. All samples were sequenced using paired-end reads with read length varying from 75 bp to 300 bp.

### Bioinformatic analysis

For bioinformatic analysis of exome sequencing data, we developed a Docker-based analysis pipeline to ease data transfer and enable the usage of each laboratory’s own computing resources. For read alignment, a faster reimplementation of the BWA mem algorithm, BWA-MEM2 (Vasimuddin et al. 2019), was used. Aligned reads were sorted and indexed using SAMtools (Li et al. 2009); duplicate read pairs were marked using the Genome Analysis ToolKit (GATK) v. 4.2.1.0 (Van der Auwera et al. 2013; DePristo et al. 2011). Next, base quality score recalibration and indel realignment was performed using GATK. Pre-processed alignments were then used for variant calling with the GATK HaplotypeCaller in the ERC GVCF mode. The source code of the pipeline can be found at https://github.com/bioinf/russian_exome_pipeline/

GVCF files were then transferred between laboratories and aggregated using the GATK GEnomicsDB engine. Aggregated GVCF files were then used for joint genotyping using GATK After joint genotyping, all sample-level genotypes with a total depth of less than 10 reads were set to missing, and variants with AC=0 were excluded. The variant callset was then annotated using the Ensembl Variant Effect Predictor (VEP) v. 104 with the RefSeq cache file for the corresponding reference genome assembly. After variant annotation, Variant Quality Score Recalibration (VQSR) filtering approach was applied; variants with truth sensitivity score between

90.0 and 99.6 (for SNPs) and 90.0 and 99.3 (for indels) were marked as medium confidence; variants with sensitivity values > 99.6 (for SNPs) and 99.3 (for indels) were excluded as low-confidence calls. The resulting multi-sample VCF file was used for variant quality control, filtering and statistical analysis (see below).

### Quality control and statistical analysis

Variant quality control and further statistical analysis of allele frequencies was performed using the BROAD Institute Hail statistical genetics library v. 0.2.63-cb767a7507c8 (https://hail.is/). First, sample-level quality control was performed using the built-in Hail functionality. Four main sample-level metrics were used: heterozygous to non-reference homozygous variant ratio (het/hom), transition-to-transversion ratio (Ti/Tv), insertion-to-deletion ratio, and mean per-sample genotype quality (GQ). The following filtering criteria were applied: 1.4 < het/hom < 2.6, 2.1 < Ti/Tv < 2.9; 0.5 < i/d < 1.0. Samples not meeting any of these criteria were removed from further analysis. We also excluded samples with less than 80% of common CDS bases covered at least 10x. In total, 621 samples were removed.

After sample-level QC, we went on to filter out related individuals. To do so, we first selected common (MAF > 5%) variants that had a high (> 99%) variant call rate across all pass-QC samples. Next, we calculated kinship statistic using the built-in Hail functionality. A maximum independent set of unrelated samples was computed using the kinship statistic value of 0.250, removing additional 330 samples.

For principal component analysis of individual genotypes, we performed HWE-normalized PCA using the built-in Hail functionality. Samples were clustered into three clusters using *k*-means clustering algorithm in the space of first 10 principal components. Allele frequencies in each cluster were determined using Hail built-in aggregation functions.

The analysis of allele frequency correlations was performed using Python v3.9 with numpy, scipy, and pandas packages. Pearson’s correlation coefficient (*r*^*2*^) was computed using a subset of variants with high (> 95%) call rate across all samples.

Variant overrepresentation was evaluated by computing a binomial *p*-value as follows:

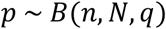

where *n* is the observed alternative allele count in the sample; *N* is the total allele number at a given variant site, *q* is the alternative allele frequency in the gnomAD non-Finnish European population, and *B* denotes the binomial distribution PDF. Only variants reported as pathogenic in ClinVar with no conflicting interpretations were used for overrepresentation testing. All variants were annotated with the inheritance pattern associated with the corresponding gene (using the OMIM catalog data) and gene-level constraint (loss-of-function variant observed to expected ratio upper fraction, LOEUF) values from gnomAD v2.1.1. Lists of variants identified as overrepresented and/or uniquely present in the dataset were manually curated to exclude misannotated variants.

### Data availability

The resulting allele frequency dataset is available through an interactive web browser at http://ruseq.ru/. To access the full VCF file with variant frequencies please contact the authors using the request form available on the website.

## Results

### Multicenter approach to the creation of Russian coding allele frequency database

To this day, the most common approach of obtaining reliable allele frequency information relies on uniform processing of genetic data generated by multiple sequencing centers and genomic laboratories. At the same time, to perform such an integration in a centralized manner requires collection of a huge amount of data, which is very intensive in terms of both time and computing resources, and also potentially requires sharing sensitive or protected information. A possible way to circumvent these difficulties is to run the identical initial steps of data analysis in a distributed manner, and then aggregate the results at the level of per-sample variant calls. Cohort variant calling methods, such as ones provided in the GATK (DePristo et al. 2011) or GLnexus (Yun et al. 2021) packages, are streamlined to process large numbers of per-sample variant calls facilitated by the usage of specialized GVCF format.

Given these developments, efficient integration of data between sequencing centres would only require a reproducible and transferable implementation of the variant calling pipeline. This goal can be met by including all the software dependencies and code into a Docker image that can then be distributed across participating laboratories (Figure 1). In this work, we have constructed such a containerized version of a bioinformatic pipeline for data analysis based on the BWA and GATK HaplotypeCaller software. During the pilot phase of the project, this pipeline was applied in two independent sequencing centers located in two major cities of the Russian Federation, Moscow (“Lab 1”) and St. Petersburg (“Lab 2” and “Lab 3”); and a total of 7,452 samples were processed in these three laboratories (4,630 - in Lab 1, and 1,737 - in Lab 2, and 1,085 - in Lab 3). Aggregation of variant calls across this set of samples allowed for more than ten-fold expansion of a previous exome-level study which included 694 samples (Barbitoff et al. 2019). The majority of samples came from patients with diagnosed monogenic disorders (5,507), while 1,945 samples were healthy donors or patient relatives, a subgroup that we extensively used for disease allele prevalence analysis described below.

**Figure 1.**
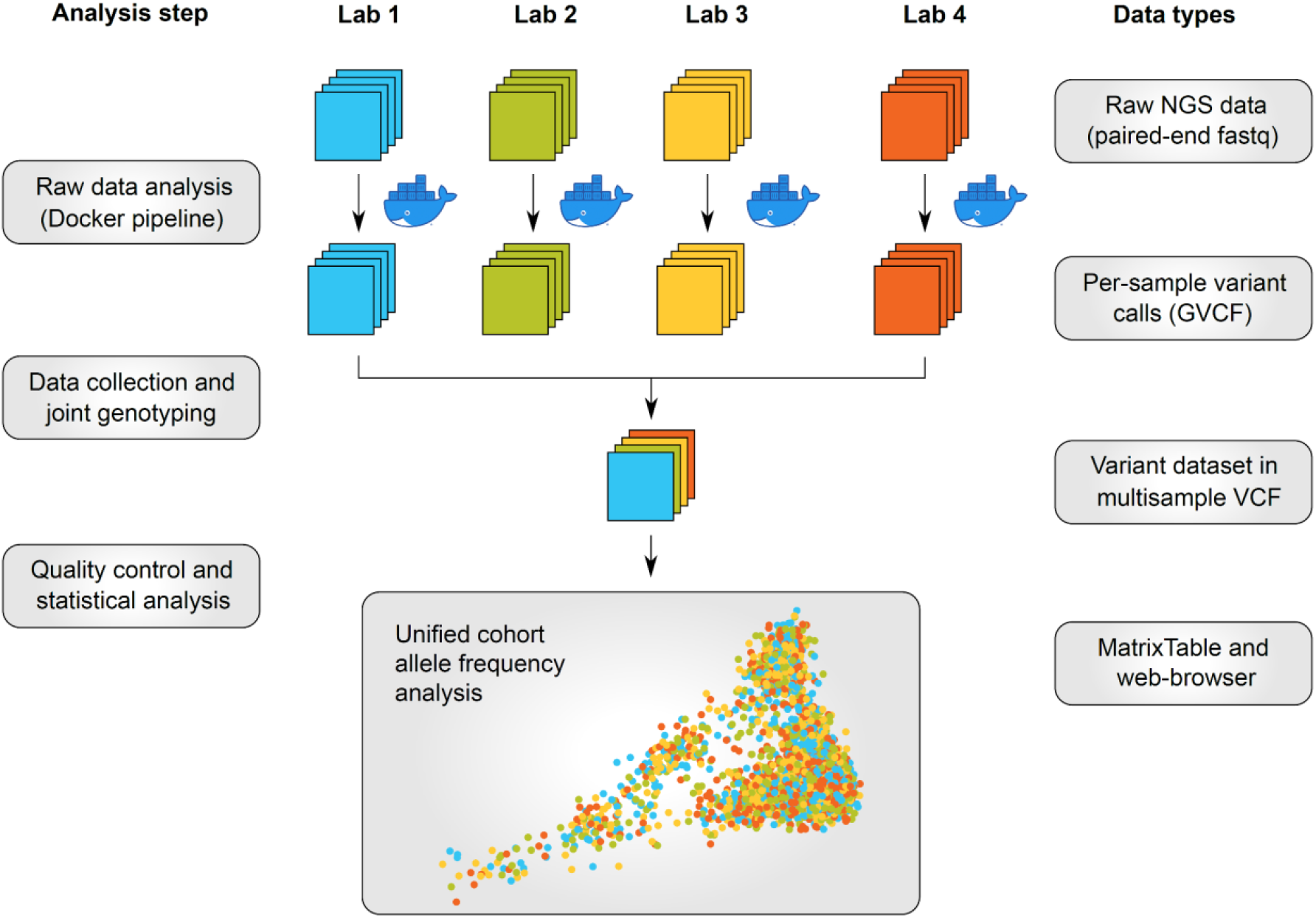
A cross-laboratory data integration strategy used for creation of an expanded reference set of allele frequencies in Russian exomes. The first steps of the data analysis, including read alignment and variant calling, are performed using a Docker-based pipeline in each laboratory, avoiding the need for raw data transfer and allowing for a distributed computing process. Variant calls in GVCF format are then aggregated, and joint genotyping is performed to obtain the final variant dataset.

Following initial data aggregation and genotyping, the dataset was subjected to extensive sample- and variant-level quality control (see Materials and Methods for more details), leaving 6,501 samples and 2,237,034 variant sites. Of these, 338,785 variant sites overlapped with the ones reported in our previous publication (Barbitoff et al. 2019). Out of all variants, 74.4 % were known (found in the latest dbSNP build), and 25.6% (572,495) were novel. In total, 1,545,345 variants (69.1%) were either non-coding or silent coding variants, 633,003 (28.3%) were missense mutations or other moderate-impact variants, and 58,686 (2.6%) variants were putative loss-of-function (pLoF) variants. Similarly to previous findings, rare and protein-damaging variants were greatly overrepresented among the novel variants. For example, only 24.9% of non-coding and silent coding variants were novel compared to as much as 43.4% of all pLoF variants. Likewise, 91.0% (2,034,859) of all variant sites were rare (MAF < 1% in the total sample) compared to as much as 99.4% (568,832) of novel variants. Finally, allele frequencies of variants in samples of each of the participating parties showed a near-perfect correlation (*r*^*2*^ = 0.999, Supplementary Figure S2).

### Analysis of the fine genetic structure of the admixed Russian population

In contrast to all previous genomic studies (Zhernakova et al. 2020; Barbitoff et al. 2019; Ramensky et al. 2021), our analysis includes a diverse set of admixed samples that can allow us to investigate the fine structure of the present-day Russian population. Indeed, principal component analysis of the individual genotypes identified several distinct clusters of samples. Presence of these clusters could not be explained by place of sample collection (Figure 2a), sequencing platform, disease status, or exome kit (Supplementary Figures S3-S5). Given this observation, we went on to group the individuals into three clusters using the unsupervised k-means algorithm. The three clusters significantly differed in size and shape. The first cluster that was dubbed “heel” was the densest and contained 5,472 (84.1%) samples. The second cluster (“ankle”) was more sparse and smaller in size, containing 729 (11.2%) samples. Finally, the third cluster (“toes”) was the smallest and the most heterogeneous, spreading over the first principal component axis (Figure 2b), and comprising 300 (4.6%) individuals.

**Figure 2.**
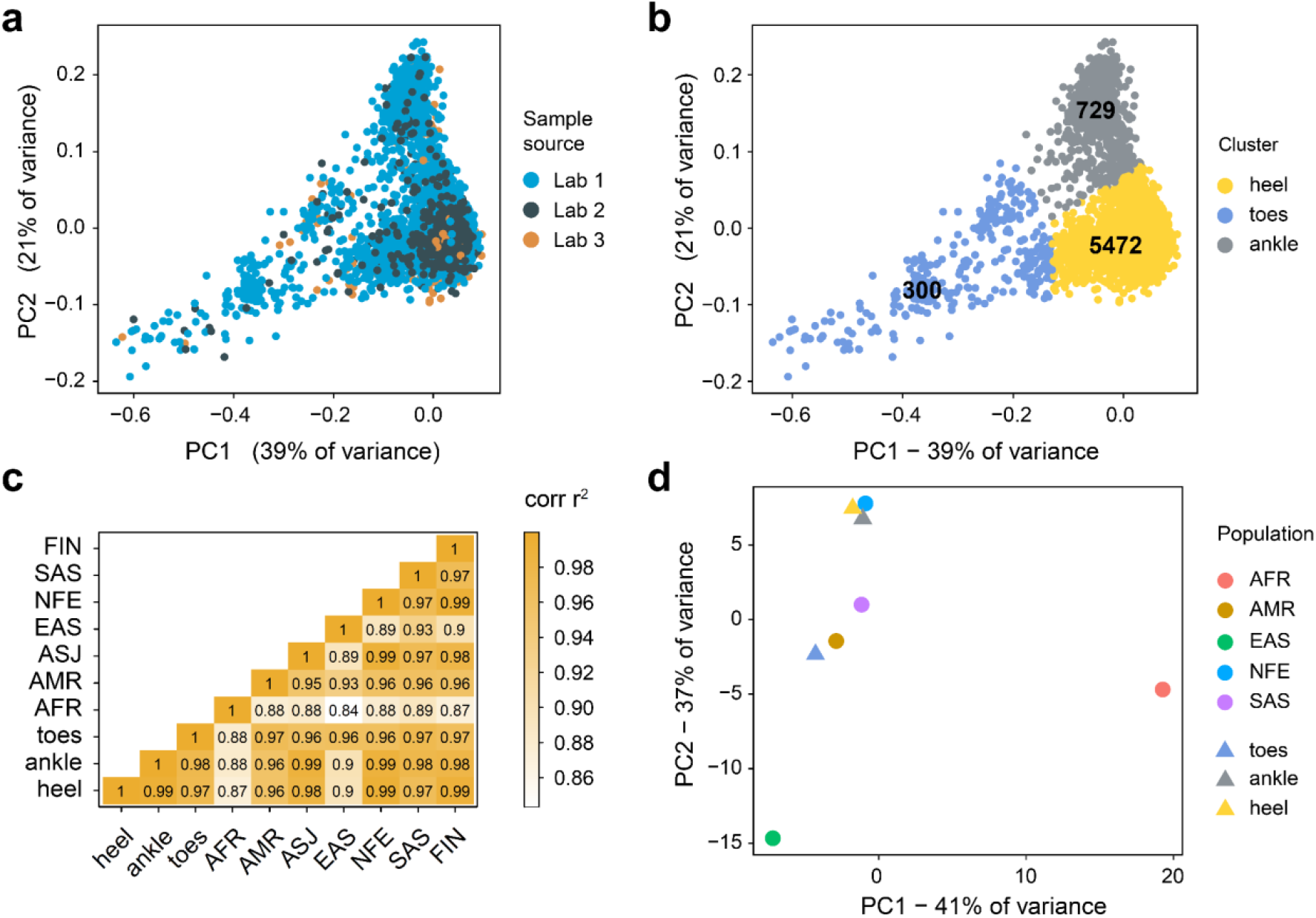
Analysis of the sub-structure of the admixed Russian population. **(a, b)** Principal component analysis of the genotypes colored by the genetic centre (a) or the results of k-means clustering in the space of first 10 principal components (b). **(c)** A heatmap showing Pearson’s correlation between common variant allele frequencies in gnomAD ancestral groups and three clusters of individuals identified by *k*-means in (b). **(d)** Principal component analysis of allele frequencies of common variants in gnomAD and three clusters of individuals identified by *k*-means in (b).

Having identified the three distinct subgroups of samples, we then asked which of the global ancestral groups are closest to these clusters. To answer this question, we first analyzed the correlation between common variant allele frequencies between each cluster and the main seven populations of gnomAD (African (AFR), Ad Mixed American (AMR), Ashkenazi Jewish (ASJ), East Asian (EAS), European - Finnish (FIN) and non-Finnish (NFE), and South Asian (SAS)). The main cluster of Russian individuals (“heel”) had the greatest correlation with the gnomAD NFE group (*r*^*2*^ = 0.992), closely followed by the Finnish population (*r*^*2*^ = 0.989). At the same time, the second (“ankle”) cluster had a high degree of AF correlation with the NFE group (*r*^*2*^ = 0.989), but was closer to Ashkenazi Jews than to the Finnish individuals from gnomAD (*r*^*2*^ = 0.988 and *r*^*2*^= 0.980, respectively), and had a much higher AF correlation with both EAS and SAS. Finally, the third (“toes”) cluster showed the greatest correlation with the SAS subpopulation from gnomAD (*r*^*2*^ = 0.974), and was equally distinct from EAS and NFE (*r*^*2*^ = 0.963 for both, Figure 2c). In addition to the observations made by the analysis of AF correlation, we performed principal component analysis of common variant allele frequencies using data from the three clusters and five major gnomAD ancestral groups (AFR, AMR, EAS, NFE, and SAS). This analysis showed that, as expected, both the “heel” and “ankle” clusters were close to the NFE group, with the “ankle” found to be closer to SAS/EAS. At the same time, the heterogeneous “toes” cluster was much more distant and appeared closer to the gnomAD SAS (Figure 2d, Supplementary Figure S2).

Given these observations, we conclude that the first cluster represents the individuals of European ancestry, *i*.*e*., native residents of the Central and Northwest Russia; the second cluster represents Southern Russia and Northern Caucasus populations, while the third cluster corresponds to patients originating from the Siberian regions and/or Asian republics of the former USSR. These assumptions were further validated by the available patient information from both sequencing centers (data not shown).

### Identification and validation of overrepresented pathogenic and benign variants

Local allele frequency reference compendia are essential for two major applications: (i) to characterize the spectrum of highly prevalent clinically significant variants that can be included into early screening programs; and (ii) to identify potentially clinically significant variants that are too common in the local population to be interpreted as disease-causing. The expanded set of allele frequencies obtained in this study may aid in solving both of these critically important tasks.

Before identifying overrepresented variants in our data, we examined allele frequencies of pathogenic variants that are known to be present at high frequency in Russia. These include (i) rs113993960 variant in *CFTR* leading to the deletion of a crucial F508 residue of the CFTR protein, and (ii) rs80338939 variant in *GJB2* linked to hearing loss and deafness (Abramov et al. 2016). In concordance with previous genetic epidemiology studies, both of these variants were present at a very high frequency in our dataset (AF = 0.0063 for rs113993960; AF = 0.0170 for rs80338939). These results validate earlier observations and show that the constructed allele frequency reference accurately captures some of the known clinically significant genetic variation in Russia.

Nearly two dozen prevalent and overrepresented pathogenic variants were reported in the two major sequencing-based studies of the Russian population (Barbitoff et al. 2019; Ramensky et al. 2021). We first questioned if the variants identified as overrepresented in earlier studies are also confirmed in our dataset. To this end, we selected variants that were reported by Barbitoff et al. and Ramensky et al. A total of 22 variants were described in these two publications (14 - in Barbitoff et al., and 10 - in Ramensky et al., with 2 overlapping variants). Of these, overrepresentation of 13 variants was successfully validated using the healthy donor subset, and of 16 - in the complete set of 6,501 samples (Supplementary Table S1).

We next went on to identify all variants that showed overrepresentation in our dataset. To this end, we computed the binomial overrepresentation p-value for all variants that were reported as pathogenic in ClinVar with no conflicting interpretations. Multiple submissions to ClinVar were required to include a variant into the analysis. As a result, 47 high-quality disease alleles were identified as overrepresented in the healthy donor subset (Table 1, Supplementary Table S2). The most common of these variants included both known high-frequency variants, such as the rs5030654 variant in *PAH* and the rs1555287300 in *ATP7B* linked to phenylketonuria and Wilson’s disease respectively, and variants that have not been previously reported as overrepresented. Notable examples of the latter category include rs554847663 in *OTOG* and rs72474224 in *GJB2* linked to autosomal recessive deafness, as well as the rs104893924 in *SLC26A2* and rs199952377 in *WDR35* causing multiple epiphyseal dysplasia and cranioectodermal dysplasia, respectively.

**Table 1.** Fifteen most common known pathogenic variants present at high frequency in RUSeq.

| Variant | Gene | gnomAD NFE AF | Allele count | RUSeq AF* | p-value | Disease |
| --- | --- | --- | --- | --- | --- | --- |
| 12:102840493G>A (rs5030858) | *PAH* | 0.15% | 23 | 0.68% | 3.56E-09 | Phenylketonuria |
| 2:151501423G>A (rs549794342) | *NEB* | 0.05% | 22 | 0.65% | 4.79E-18 | Nemaline myopathy |
| 13:51944145G>T (rs76151636) | *ATP7B* | 0.13% | 19 | 0.57% | 1.53E-07 | Wilson disease |
| 13:113118668C>T (rs41286844) | *C8B* | 0.19% | 18 | 0.53% | 1.11E-04 | C8 deficiency, type II |
| 1:56940965G>A (rs36209567) | *F7* | 0.10% | 18 | 0.54% | 3.09E-08 | Factor VII deficiency |
| 11:17574890C>T (rs554847663) | *OTOG* | 0.08% | 17 | 0.53% | 1.14E-09 | Deafness |
| 11:71441401C>T (rs11555217) | *DHCR7* | 0.14% | 17 | 0.50% | 1.21E-05 | Smith-Lemli-Opitz syndrome |
| 10:89222511C>T (rs116928232) | *LIPA* | 0.13% | 15 | 0.46% | 3.39E-05 | Cholesteryl ester storage disease |
| 13:20189473C>T (rs72474224) | *GJB2* | 0.13% | 15 | 0.45% | 5.74E-05 | Deafness |
| 11:89178603G>A (rs61754365) | *TYR* | 0.03% | 14 | 0.42% | 1.34E-11 | Albinism, oculocutaneous |
| 16:3243310A>G (rs28940579) | *MEFV* | 0.09% | 12 | 0.35% | 1.10E-04 | Familial Mediterranean fever |
| 20:54158136G>A (rs114368325) | *CYP24A1* | 0.11% | 12 | 0.36% | 3.82E-04 | Hypercalcemia, infantile |
| 5:149981550T>A (rs104893924) | *SLC26A2* | 0.02% | 11 | 0.33% | 1.05E-09 | Multiple epiphyseal dysplasia |
| 2:19941796A>C (rs199952377) | *WDR35* | 0.03% | 10 | 0.31% | 1.03E-07 | Cranioectodermal dysplasia |
| 6:80201023G>A (rs386834233) | *BCKDHB* | 0.04% | 10 | 0.30% | 1.15E-06 | Maple syrup urine disease |

\* - AF is given with reference to the healthy donor subgroup.

Notably, several rare variants showed extremely high levels of fold enrichment in Russia. For example, the rs376910645 variant in the *ATP7B* gene was observed 3 times in the subset of healthy Russian donors (AF = 0.0009). The frequency of this variant in the gnomAD non-Finnish European population is approximately 100 times lower, with only one observation across all gnomAD r. 2.1 exome and gnomAD r. 3.1 genome samples. Another example of a highly enriched pathogenic variant is the rs749076525 in the *DHCR7* gene encoding sterol delta-7-reductase (RUSeq AF = 0.0012, fold enrichment versus gnomAD NFE = 67.4). Mutations in this gene cause Smith-Lemli-Opitz syndrome, an autosomal-recessive condition which involves multiple congenital malformations and intellectual disability. Remarkably, one more variant in *DHCR7* was also found to be significantly overrepresented (rs11555217, observed 17 times in RUSeq healthy donor group). Similarly, two factor VII deficiency-causing variants were found to be overrepresented in the *F7* gene (rs36209567 and rs121964931, RUSeq AF = 0.0054 and 0.0009, respectively), with the latter variant also having a 25-fold enrichment in the Russian population. Taken together, these data suggest that both Smith-Lemli-Opitz syndrome and factor VII deficiency should have high incidence in the Russian population. For the *F7* gene, similar conclusions were made by us earlier using a smaller set of WES samples (Barbitoff et al. 2019).

Having characterized the spectrum of overrepresented pathogenic alleles in our dataset, we then went on to identify potentially clinically significant variants that are present in healthy patients but are not found in gnomAD. We began by identifying known pathogenic variants missing from gnomAD v2.1.1 data that are reported in the ClinVar database. In total, we discovered 173 such variants, with 53 of them located in genes with autosomal-dominant disease inheritance, and 44 being high-confidence variants with multiple submitters (Supplementary Table S3). In addition to these variants, we also searched for potentially clinically significant variants that are absent from gnomAD but are present in the healthy subgroup. In total, more than 100 putative loss-of-function (pLoF) variants present in healthy patients were identified. Of these, 59 variants localized in genes with high degree of evolutionary conservation according to gnomAD-derived metrics (pLI, LOEUF) and with known connection to autosomal dominant phenotypes. A total of 46 variants passed the additional manual curation step. Selected variants are listed in Supplementary Table S3. Among the known and expected pathogenic variants present in our sample, the most notable example is the rs1064793825 variant in *MSH2* possibly causing hereditary colorectal cancer. A carrier subject has not yet displayed any symptoms of the disorder.

However, family history of the carrier listed several cases of cancer, suggesting that the disease is likely to manifest in the near future. Furthermore, we discovered 2 known pathogenic variants that were present in both healthy and diseased donor subgroups. The first variant, rs786205232 (1:110603893C>T), is a missense variant in the *KCNA2* gene responsible for developmental and epileptic encephalopathy 32 (MIM 616366). The disease allele was observed twice in the diseased cohort and once among the healthy donors. A similar distribution was observed for the other variant, rs59285727 (17:44915251C>T) in *GFAP*, linked to the juvenile form of the Alexander disease. The latter example is especially noteworthy as the identified mutation is commonly observed in individuals with the Alexander disease (Srivastava, Waldman, and Naidu 2002).

Taken together, we described two important categories of genetic variants in our dataset: (i) known pathogenic variants that are present at higher frequency in Russia, and (ii) variants with presumed pathogenic effects that are too common in Russia and/or are present in the subgroup of healthy individuals. Both of these groups of variants can be used in NGS-based assays in Russia to assist variant interpretation in clinical practice. All of the variant frequency data presented here are available to the community of medical geneticists through a web-based variant portal at http://ruseq.ru.

## Discussion

As shown in multiple studies, population-adjusted allele frequency information is useful for efficient filtering of candidate pathogenic variants. For example, filtering of variants by maximum allele frequency across populations rather than global allele frequency helps to decrease the number of candidate pathogenic variants per individual genome (Lek et al. 2016). Despite the great availability of global allele frequency information in such resources as gnomAD, many nations and local populations remain poorly represented from a genetic perspective. Russia, being the world’s largest country by total land area, has long remained the widest blank spot on the genetic map of the world (Oleksyk, Brukhin, and O’Brien 2015).

Three major studies have addressed the problem of limited genomic information about the Russian population. One of the studies included only healthy donors from different ethnic backgrounds (Zhernakova et al. 2020); however, this study suffered from a very low sample size that was not sufficient for clinical purposes. An earlier study from our group included 694 samples and provided the first exome-wide estimates of disease allele prevalence (Barbitoff et al. 2019). Finally, a very recent study by Ramensky et al. included 1,658 healthy controls; unfortunately, this study was based on a targeted sequencing of a small number of genes (242) (Ramensky et al., 2021). Despite the aforementioned limitations, data obtained in previous analyses have already been used for interpretation of clinical significance of variants (Glotov et al. 2019; Maksimov et al. 2020) and population genetics analyses (e.g., (Shikov et al. 2020; Pinheiro, Sperb-Ludwig, and Schwartz 2021)). Exome-based allele frequencies have been included into several databases, including a database of *BRCA1/BRCA2* gene mutations (https://oncobrca.ru/). A dramatic (more than tenfold, 7,452 compared to 694) increase in the sample size achieved in this study brings the available allele frequency reference for the Russian population to the level of such a project as ESP6500 (Fu et al. 2013), greatly aiding clinical specialists in variant interpretation in NGS-based assays. Increase in the sample size allowed us to identify genetically distinct clusters in the admixed sample of Russian residents, which were in line with earlier studies (Zhernakova et al. 2020). At the same time, existence of distinct clusters imply that different genetic disease risk factors are present in different geographical regions of Russia, which is well known to the Russian medical genetic community. This finding comes as no surprise, but predicates the need for further expansion and diversification of the dataset.

As described in the previous section, we have successfully validated some of the earlier observations regarding overrepresentation of disease alleles in the Russian population. Out of 22 disease-associated variants that were identified in (Barbitoff et al. 2019; Ramensky et al. 2021) we confirmed overrepresentation of 18 variants (81.8%), and identified many novel pathogenic alleles that have greater incidence in the Russian Federation compared to other European populations (gnomAD NFE group). For several of the identified variants, their increased prevalence in Russia has been noted in gene-level studies (Abramov et al. 2015; Balashova et al. 2020). For other variants, such as the rs554847663 variant in *OTOG*, no previous reports were published. Hence, these variants represent novel candidates that are worth looking into in further genetic epidemiology studies. In addition to the overrepresented variants, we find a limited set of variants in AD genes that are annotated as pathogenic in ClinVar and linked to severe disorders but are present in healthy control individuals in Russia (Supplementary Table S2). This information is important for clinical interpretation of such variants, especially in the Russian population.

While the current sample size allows for more unbiased conclusions regarding the genetic structure of the Russian population, we can still expect a large number of rare genetic variants in the rest of the population that were not covered by our analysis. Recent studies have shown that mutational saturation can only be achieved by integrating hundreds of thousands of samples (Agarwal and Przeworski 2021). Hence, further aggregation of data from sequencing centers across Russia, sequencing of more healthy donors, and inclusion of patients from distinct regions (such as in the initial design of the Genomes Russia project (Oleksyk, Brukhin, and O’Brien 2015)) are all required to fully characterize the genetic variation spectrum of present-day Russia.

## Data Availability

Variant frequency level data produced in the present study are available via form submission available at ruseq.ru.

http://ruseq.ru

## Acknowledgements

We want to thank all the patients who agreed to provide their data for population analysis. We also thank Mrinal Vashisth and Mary K. Futey for their help with development of the data analysis pipeline. Statistical analysis and expert review of the results was supported by the Systems Biology Fellowship to Y.A.B. the Presidential Fellowship for Young Scientists (grant no. SP-4503.2021.4) to Y.A.B., and D.O. Ott Research Institute of Obstetrics, Gynaecology and Reproductology, project 558-2019-0012 (AAAA-A19119021290033-1) of FSBSI.

## References

Abramov, DD, MV Belousova, VV Kadochnikova, AA Ragimov, and D Yu Trofimov. 2016. “Carrier Frequency of GJB2 and GALT Mutations Associated with Sensorineural Hearing Loss and Galactosemia in the Russian Population.” Bulletin of Russian State Medical University, no. 6.

Abramov, DD, VV Kadochnikova, EG Yakimova, MV Belousova, AV Maerle, IV Sergeev, AA Ragimov, AE Donnikov, and DY Trofimov. 2015. “High Carrier Frequency of CFTR Gene Mutations Associated with Cystic Fibrosis, and PAH Gene Mutations Associated with Phenylketonuria in Russian Population.” Bulletin of RSMU 4: 32–35.

Agarwal, Ipsita, and Molly Przeworski. 2021. “Mutation Saturation for Fitness Effects at Human CpG Sites.” Preprint. Evolutionary Biology. https://doi.org/10.1101/2021.06.02.446661.

Auton, Adam, Gonçalo R. Abecasis, Steering committee, David M. Altshuler, Richard M. Durbin, Gonçalo R. Abecasis, David R. Bentley, et al. 2015. “A Global Reference for Human Genetic Variation.” Nature 526 (7571): 68–74. https://doi.org/10.1038/nature15393.

Balashova, Mariya S., Inna G. Tuluzanovskaya, Oleg S. Glotov, Andrey S. Glotov, Yury A. Barbitoff, Mikhail A. Fedyakov, Diana A. Alaverdian, et al. 2020. “The Spectrum of Pathogenic Variants of the ATP7B Gene in Wilson Disease in the Russian Federation.” Journal of Trace Elements in Medicine and Biology 59 (May): 126420. https://doi.org/10.1016/j.jtemb.2019.126420.

Barbitoff, Yury A., Dmitrii E. Polev, Andrey S. Glotov, Elena A. Serebryakova, Irina V. Shcherbakova, Artem M. Kiselev, Anna A. Kostareva, Oleg S. Glotov, and Alexander V. Predeus. 2020. “Systematic Dissection of Biases in Whole-Exome and Whole-Genome Sequencing Reveals Major Determinants of Coding Sequence Coverage.” Scientific Reports 10 (1): 2057. https://doi.org/10.1038/s41598-020-59026-y.

Barbitoff, Yury A., Rostislav K. Skitchenko, Olga I. Poleshchuk, Anton E. Shikov, Elena A. Serebryakova, Yulia A. Nasykhova, Dmitrii E. Polev, et al. 2019. “Whole-exome Sequencing Provides Insights into Monogenic Disease Prevalence in Northwest Russia.” Molecular Genetics & Genomic Medicine 7 (11). https://doi.org/10.1002/mgg3.964.

Bick, Alexander G., Joshua S. Weinstock, Satish K. Nandakumar, Charles P. Fulco, Erik L. Bao, Seyedeh M. Zekavat, Mindy D. Szeto, et al. 2020. “Inherited Causes of Clonal Haematopoiesis in 97,691 Whole Genomes.” Nature 586 (7831): 763–68. https://doi.org/10.1038/s41586-020-2819-2.

Biesecker, Leslie G., and Robert C. Green. 2014. “Diagnostic Clinical Genome and Exome Sequencing.” New England Journal of Medicine 370 (25): 2418–25. https://doi.org/10.1056/NEJMra1312543.

Boomsma, Dorret I, Cisca Wijmenga, Eline P Slagboom, Morris A Swertz, Lennart C Karssen, Abdel Abdellaoui, Kai Ye, et al. 2014. “The Genome of the Netherlands: Design, and Project Goals.” European Journal of Human Genetics 22 (2): 221–27. https://doi.org/10.1038/ejhg.2013.118.

DePristo, Mark A, Eric Banks, Ryan Poplin, Kiran V Garimella, Jared R Maguire, Christopher Hartl, Anthony A Philippakis, et al. 2011. “A Framework for Variation Discovery and Genotyping Using Next-Generation DNA Sequencing Data.” Nature Genetics 43 (5): 491– 98. https://doi.org/10.1038/ng.806.

Fu, Wenqing, Timothy D. O’Connor, Goo Jun, Hyun Min Kang, Goncalo Abecasis, Suzanne M. Leal, Stacey Gabriel, et al. 2013. “Analysis of 6,515 Exomes Reveals the Recent Origin of Most Human Protein-Coding Variants.” Nature 493 (7431): 216–20. https://doi.org/10.1038/nature11690.

Gao, Yang, Chao Zhang, Liyun Yuan, YunChao Ling, Xiaoji Wang, Chang Liu, Yuwen Pan, et al. 2020. “PGG.Han: The Han Chinese Genome Database and Analysis Platform.” Nucleic Acids Research 48 (D1): D971–76. https://doi.org/10.1093/nar/gkz829.

Glotov, Oleg, Elena Serebryakova, Mariia Turkunova, Olga Efimova, Andrey Glotov, Yury Barbitoff, Yulia Nasykhova, et al. 2019. “Whole-exome Sequencing in Russian Children with Non-type 1 Diabetes Mellitus Reveals a Wide Spectrum of Genetic Variants in MODY-related and Unrelated Genes.” Molecular Medicine Reports, October. https://doi.org/10.3892/mmr.2019.10751.

Karczewski, Konrad J., Laurent C. Francioli, Grace Tiao, Beryl B. Cummings, Jessica Alföldi, Qingbo Wang, Ryan L. Collins, et al. 2020. “The Mutational Constraint Spectrum Quantified from Variation in 141,456 Humans.” Nature 581 (7809): 434–43. https://doi.org/10.1038/s41586-020-2308-7.

Lek, Monkol, Konrad J. Karczewski, Eric V. Minikel, Kaitlin E. Samocha, Eric Banks, Timothy Fennell, Anne H. O’Donnell-Luria, et al. 2016. “Analysis of Protein-Coding Genetic Variation in 60,706 Humans.” Nature 536 (7616): 285–91. https://doi.org/10.1038/nature19057.

Li, H., B. Handsaker, A. Wysoker, T. Fennell, J. Ruan, N. Homer, G. Marth, G. Abecasis, R. Durbin, and 1000 Genome Project Data Processing Subgroup. 2009. “The Sequence Alignment/Map Format and SAMtools.” Bioinformatics 25 (16): 2078–79. https://doi.org/10.1093/bioinformatics/btp352.

Maksimov, V. N., D. E. Ivanoshchuk, P. S. Orlov, A. A. Ivanova, S. K. Malyutina, S. V. Maksimova, I. A. Rodina, O. V. Khamovich, V. P. Novoselov, and M. I. Voevoda. 2020. “Next Generation Sequencing in Sudden Cardiac Death (Pilot Study).” Russian Journal of Cardiology 25 (10): 3880. https://doi.org/10.15829/1560-4071-2020-3880.

Martin, Alicia R, Solomon Teferra, Marlo Möller, Eileen G Hoal, and Mark J Daly. 2018. “The Critical Needs and Challenges for Genetic Architecture Studies in Africa.” Current Opinion in Genetics & Development 53 (December): 113–20. https://doi.org/10.1016/j.gde.2018.08.005.

Nykamp, Keith, Michael Anderson, Martin Powers, John Garcia, Blanca Herrera, Yuan-Yuan Ho, Yuya Kobayashi, et al. 2017. “Sherloc: A Comprehensive Refinement of the ACMG–AMP Variant Classification Criteria.” Genetics in Medicine 19 (10): 1105–17. https://doi.org/10.1038/gim.2017.37.

Oleksyk, Taras K., Vladimir Brukhin, and Stephen J. O’Brien. 2015. “The Genome Russia Project: Closing the Largest Remaining Omission on the World Genome Map.” GigaScience 4 (1): 53. https://doi.org/10.1186/s13742-015-0095-0.

Pinheiro, Franciele C., Fernanda Sperb-Ludwig, and Ida V. D. Schwartz. 2021. “Epidemiological Aspects of Hereditary Fructose Intolerance: A Database Study.” Human Mutation, September, humu.24282. https://doi.org/10.1002/humu.24282.

Ramensky, Vasily E., Alexandra I. Ershova, Marija Zaicenoka, Anna V. Kiseleva, Anastasia A. Zharikova, Yuri V. Vyatkin, Evgeniia A. Sotnikova, et al. 2021. “Targeted Sequencing of 242 Clinically Important Genes in the Russian Population From the Ivanovo Region.” Frontiers in Genetics 12 (October): 709419. https://doi.org/10.3389/fgene.2021.709419.

Richards, Sue, Nazneen Aziz, Sherri Bale, David Bick, Soma Das, Julie Gastier-Foster, Wayne W. Grody, et al. 2015. “Standards and Guidelines for the Interpretation of Sequence Variants: A Joint Consensus Recommendation of the American College of Medical Genetics and Genomics and the Association for Molecular Pathology.” Genetics in Medicine 17 (5): 405–23. https://doi.org/10.1038/gim.2015.30.

Ryzhkova, OP, OL Kardymon, EB Prohorchuk, FA Konovalov, AB Maslennikov, VA Stepanov, AA Afanasyev, et al. 2018. “Guidelines for the interpretation of massive parallel sequencing variants (update 2018, v2).” Meditsinskaia genetika, no. 2.

Shikov, Anton E., Rostislav K. Skitchenko, Alexander V. Predeus, and Yury A. Barbitoff. 2020. “Phenome-Wide Functional Dissection of Pleiotropic Effects Highlights Key Molecular Pathways for Human Complex Traits.” Scientific Reports 10 (1): 1037. https://doi.org/10.1038/s41598-020-58040-4.

Srivastava, S, A Waldman, and S Naidu. 2002. Alexander Disease. 2002 Nov 15 [Updated 2020 Nov 12]. In: Adam MP, Everman DB, Mirzaa GM, et al., Editors. GeneReviews® [Internet]. Seattle (WA): University of Washington, Seattle; 1993–2022. Available From: https://www.ncbi.nlm.nih.gov/books/NBK1172/.

Van der Auwera, Geraldine A., Mauricio O. Carneiro, Christopher Hartl, Ryan Poplin, Guillermo del Angel, Ami Levy-Moonshine, Tadeusz Jordan, et al. 2013. “From FastQ Data to High-Confidence Variant Calls: The Genome Analysis Toolkit Best Practices Pipeline: The Genome Analysis Toolkit Best Practices Pipeline.” In Current Protocols in Bioinformatics, edited by Alex Bateman, William R. Pearson, Lincoln D. Stein, Gary D. Stormo, and John R. Yates, 11.10.1-11.10.33. Hoboken, NJ, USA: John Wiley & Sons, Inc. https://doi.org/10.1002/0471250953.bi1110s43.

Vasimuddin, Md., Sanchit Misra, Heng Li, and Srinivas Aluru. 2019. “Efficient Architecture-Aware Acceleration of BWA-MEM for Multicore Systems.” In 2019 IEEE International Parallel and Distributed Processing Symposium (IPDPS), 314–24. Rio de Janeiro, Brazil: IEEE. https://doi.org/10.1109/IPDPS.2019.00041.

Wong, Karen H. Y., Walfred Ma, Chun-Yu Wei, Erh-Chan Yeh, Wan-Jia Lin, Elin H. F. Wang, Jen-Ping Su, et al. 2020. “Towards a Reference Genome That Captures Global Genetic Diversity.” Nature Communications 11 (1): 5482. https://doi.org/10.1038/s41467-020-19311-w.

Wright, Caroline F., David R. FitzPatrick, and Helen V. Firth. 2018. “Paediatric Genomics: Diagnosing Rare Disease in Children.” Nature Reviews Genetics 19 (5): 253–68. https://doi.org/10.1038/nrg.2017.116.

Yun, Taedong, Helen Li, Pi-Chuan Chang, Michael F Lin, Andrew Carroll, and Cory Y McLean. 2021. “Accurate, Scalable Cohort Variant Calls Using DeepVariant and GLnexus.” Edited by Peter Robinson. Bioinformatics 36 (24): 5582–89. https://doi.org/10.1093/bioinformatics/btaa1081.

Zhernakova, Daria V., Vladimir Brukhin, Sergey Malov, Taras K. Oleksyk, Klaus Peter Koepfli, Anna Zhuk, Pavel Dobrynin, et al. 2020. “Genome-Wide Sequence Analyses of Ethnic Populations across Russia.” Genomics 112 (1): 442–58. https://doi.org/10.1016/j.ygeno.2019.03.007.

