## Supplementary Figures S1-S5 for "Expanding the Russian allele frequency reference via cross-laboratory data integration: insights from 7,452 exome samples"

### Supplementary Information

### Supplementary Figures

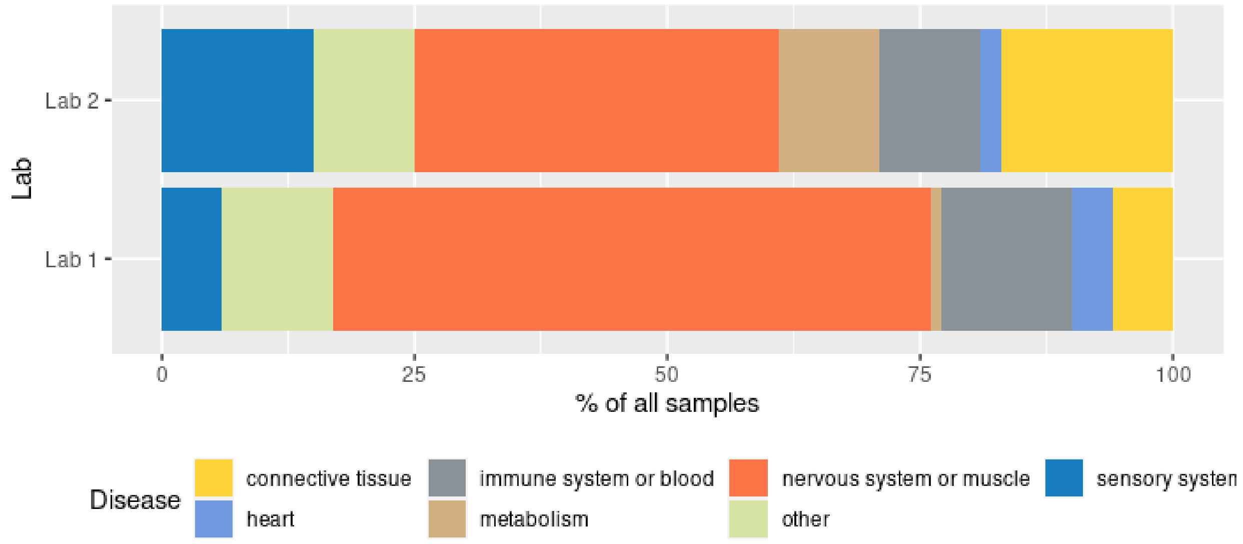


**Figure S1.** Approximate distribution of disease groups in the subset of diseased individuals included into the analysis. For Lab 3, most of the samples come from healthy donors. For the rest of the dataset, nosology data are not available.

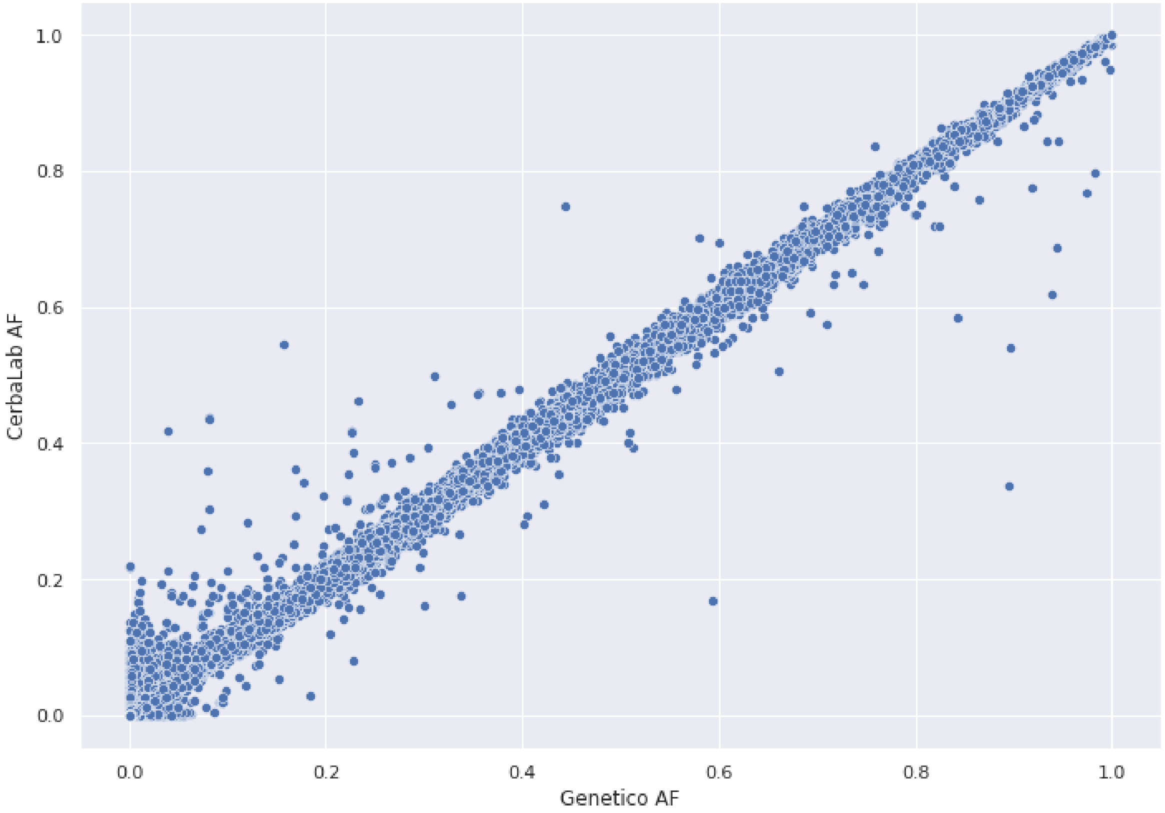


**Figure S2.** A scatterplot showing the correspondence between alternative allele frequencies for variants with call rate greater than 80% in samples from both participating laboratories. Pearson's $r^2$ for the frequencies is 0.999.

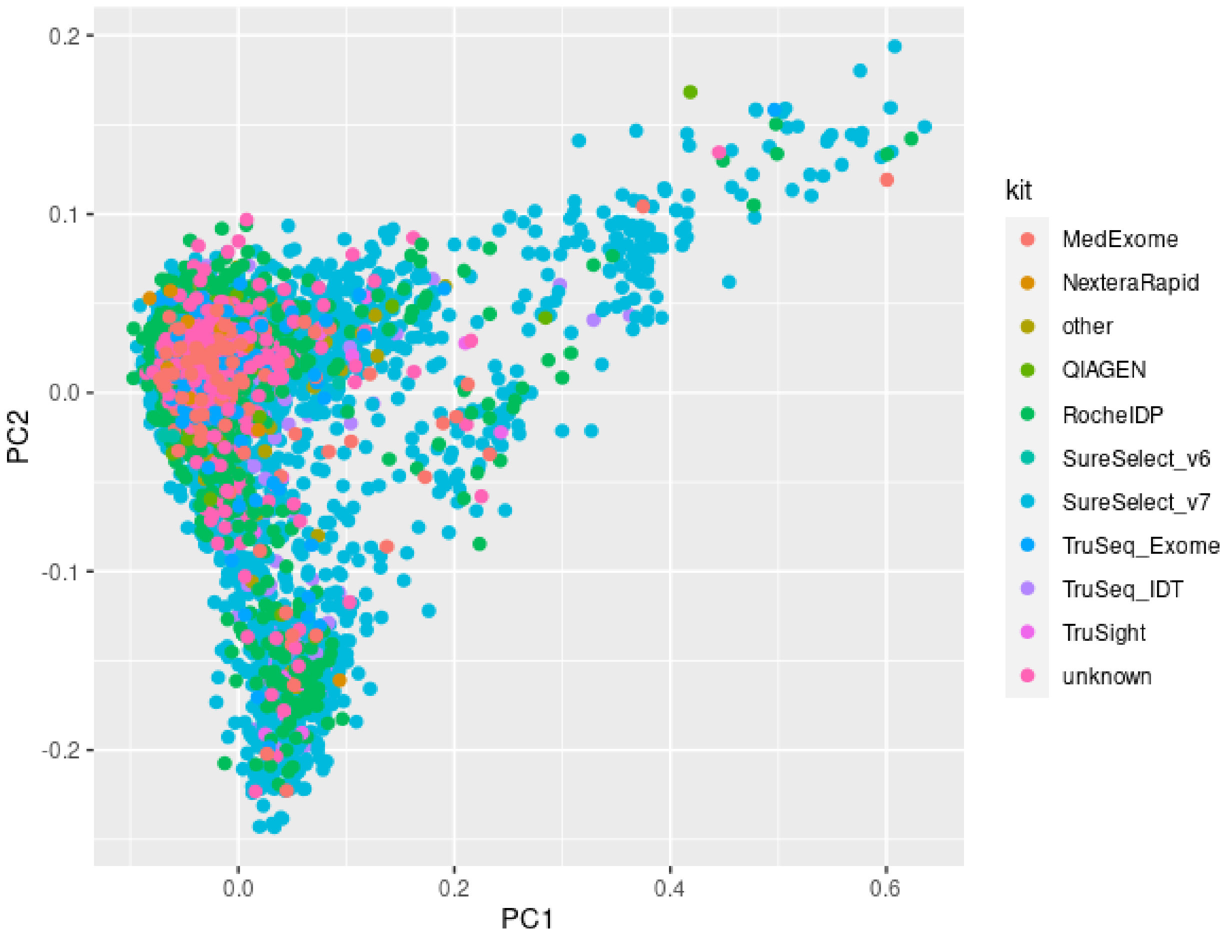


**Figure S3.** A scatterplot showing the results of principal component analysis of individual genotypes coloured by capture probes used for library preparation (A).

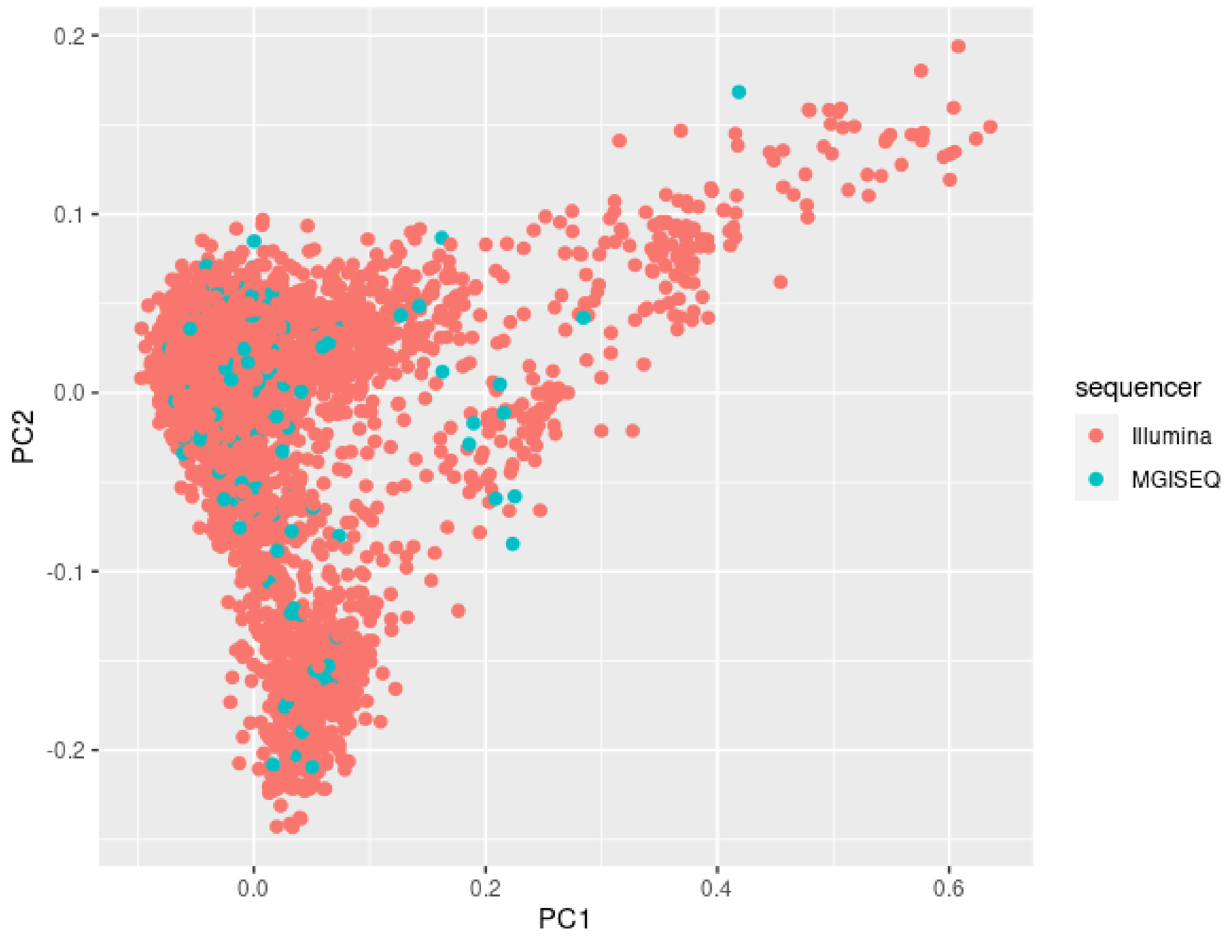


**Figure S4.** A scatterplot showing the results of principal component analysis of individual genotypes coloured by sequencing machine used.

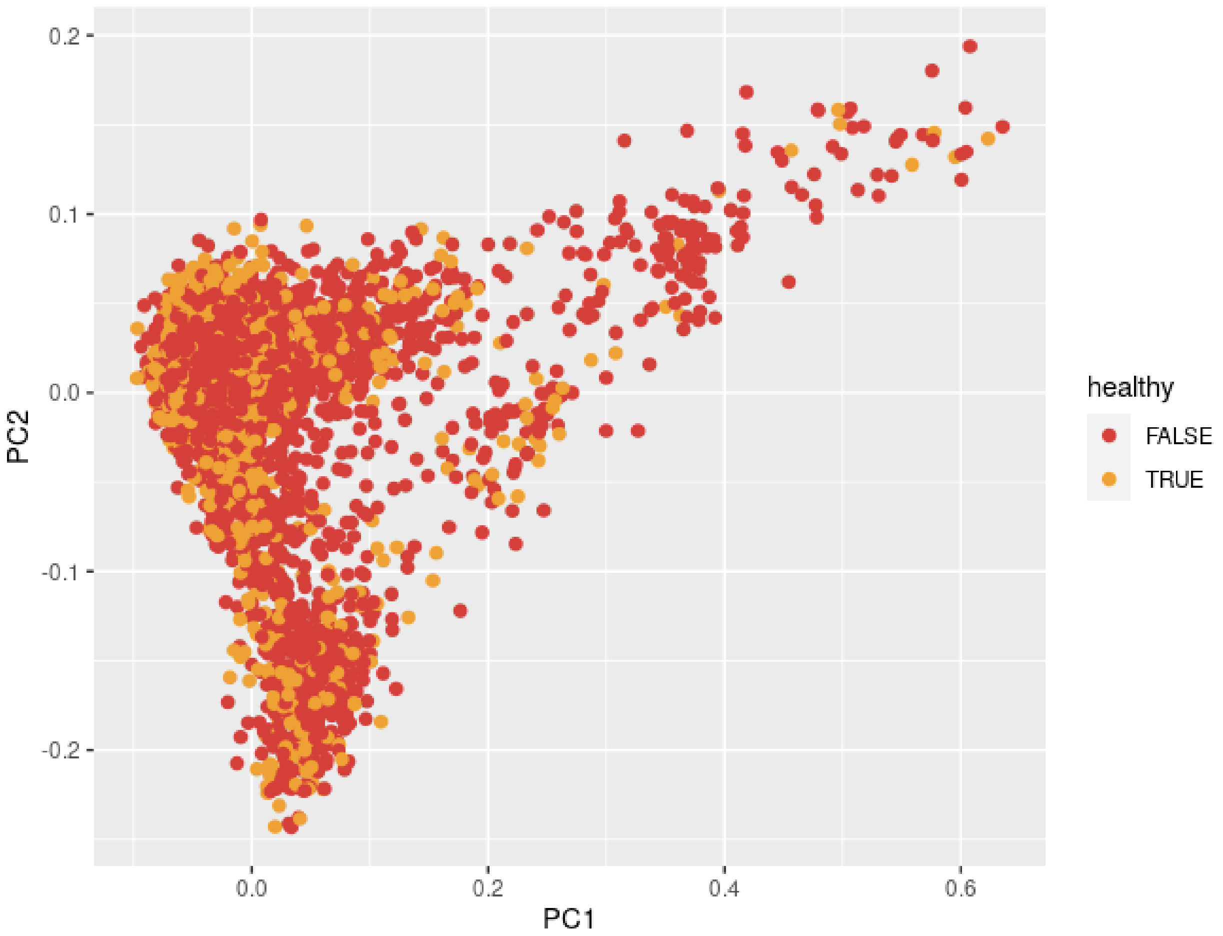


**Figure S5.** A scatterplot showing the results of principal component analysis of individual genotypes coloured by disease status.
